# A third COVID-19 vaccine shot markedly boosts neutralizing antibody potency and breadth

**DOI:** 10.1101/2021.08.11.21261670

**Authors:** Sho Iketani, Lihong Liu, Manoj S. Nair, Hiroshi Mohri, Maple Wang, Yaoxing Huang, David D. Ho

## Abstract

COVID-19 (coronavirus disease 2019) vaccines have been rapidly developed and deployed globally as a measure to combat the disease. These vaccines have been demonstrated to confer significant protection, but there have been reports of temporal decay in antibody titer. Furthermore, several variants have been identified with variable degrees of antibody resistance. These two factors suggest that a booster vaccination may be worthy of consideration. While such a booster dose has been studied as a series of three homologous vaccines in healthy individuals, to our knowledge, information on a heterologous regimen remains unreported, despite the practical benefits of such a scheme. Here, in this observational study, we investigated the serological profile of four healthy individuals who received two doses of the BNT162b2 vaccine, followed by a third booster dose with the Ad26.COV2.S vaccine. We found that while all individuals had spike-binding antibodies at each of the timepoints tested, there was an appreciable drop in titer by four months following the second vaccination. The third vaccine dose robustly increased titers beyond that of two vaccinations, and these elicited antibodies had neutralizing capability against all SARS-CoV-2 strains tested in both a recombinant vesicular stomatitis virus-based pseudovirus assay and an authentic SARS-CoV-2 assay, except for one individual against B.1.351 in the latter assay. Thus, a third COVID-19 vaccine dose in healthy individuals promoted not just neutralizing antibody potency, but also induced breadth against dominant SARS-CoV-2 variants.

**Significance:** COVID-19 vaccines confer protection from symptomatic disease, but the elicited antibody titer has been found to decrease with time. Furthermore, SARS-CoV-2 variants with relative resistance against antibody neutralization have been identified. To overcome such issues, a third vaccine dose applied as a booster vaccine may be necessary. We studied four healthy individuals who received a heterologous booster dose as a third vaccine. All of these individuals had heightened neutralizing antibody titer following the booster vaccination, and could neutralize nearly all variants tested. Thus, a heterologous third COVID-19 vaccine dose may be a mechanism to both heighten and broaden antibody titers, and could be an additional strategy for controlling the SARS-CoV-2 pandemic.

## Introduction

Multiple vaccines against SARS-CoV-2 (severe acute respiratory syndrome coronavirus 2) have been demonstrated to protect against COVID-19 (coronavirus disease 2019) (1, 2). While these vaccines have been promising in reducing symptomatic infections, the decay of neutralizing antibodies has been documented following vaccination (3, 4). Such decay is particularly pertinent due to the emergence of SARS-CoV-2 variants with relative resistance to vaccine-elicited antibodies (5-7). Consequently, losses in vaccine-mediated protection against select variants have been observed, and breakthrough infections in vaccinated individuals have been reported (8, 9). These studies suggest that a booster vaccine may be warranted. Such a concept has been studied in organ transplant patients (10, 11) and in homologous three-dose vaccination courses in healthy individuals (12), but to our knowledge, not as heterologous three-dose vaccinations in healthy individuals. Such a heterologous vaccination regimen has many practical benefits, as emphasized by studies on two heterologous vaccine schedules (13, 14). Here, we report on the antibody responses of four healthy individuals who received a two-dose course of BNT162b2 vaccination (mRNA-based vaccine, Pfizer-BioNTech), and then subsequently received the Ad26.COV2.S vaccine (adenovirus vector-based vaccine, Johnson & Johnson) as a third vaccination.

## Results and Discussion

We first confirmed that the four individuals in this study did not have anti-nucleoprotein antibodies, suggesting that their responses reflected only the immunogenicity of the vaccines (**Figure 1A**). All four individuals had spike-binding antibodies at all timepoints tested, both against the non-variant strain and B.1.351, but had demonstrable loss in binding titer by four months following their second vaccination. Robust increases in binding titer were observed following a third vaccination, greater than that achieved after two vaccine doses (**Figure 1B**). These elicited antibodies were found to have neutralizing capability against all variant pseudoviruses, as well as all authentic SARS-CoV-2 strains tested except for the case of Volunteer #4 against B.1.351 (**Figure 2**). The highest titers were observed against the non-variant strain and B.1.1.7. The increases in plasma neutralization titers (ID_50_) ranged from 10.9 to 21.1-fold in the pseudovirus neutralization assay and 14.8 to 32.4-fold in the authentic virus neutralization assay. One individual, Volunteer #1, even had heightened neutralizing titer against SARS-CoV.

**Figure 1.**
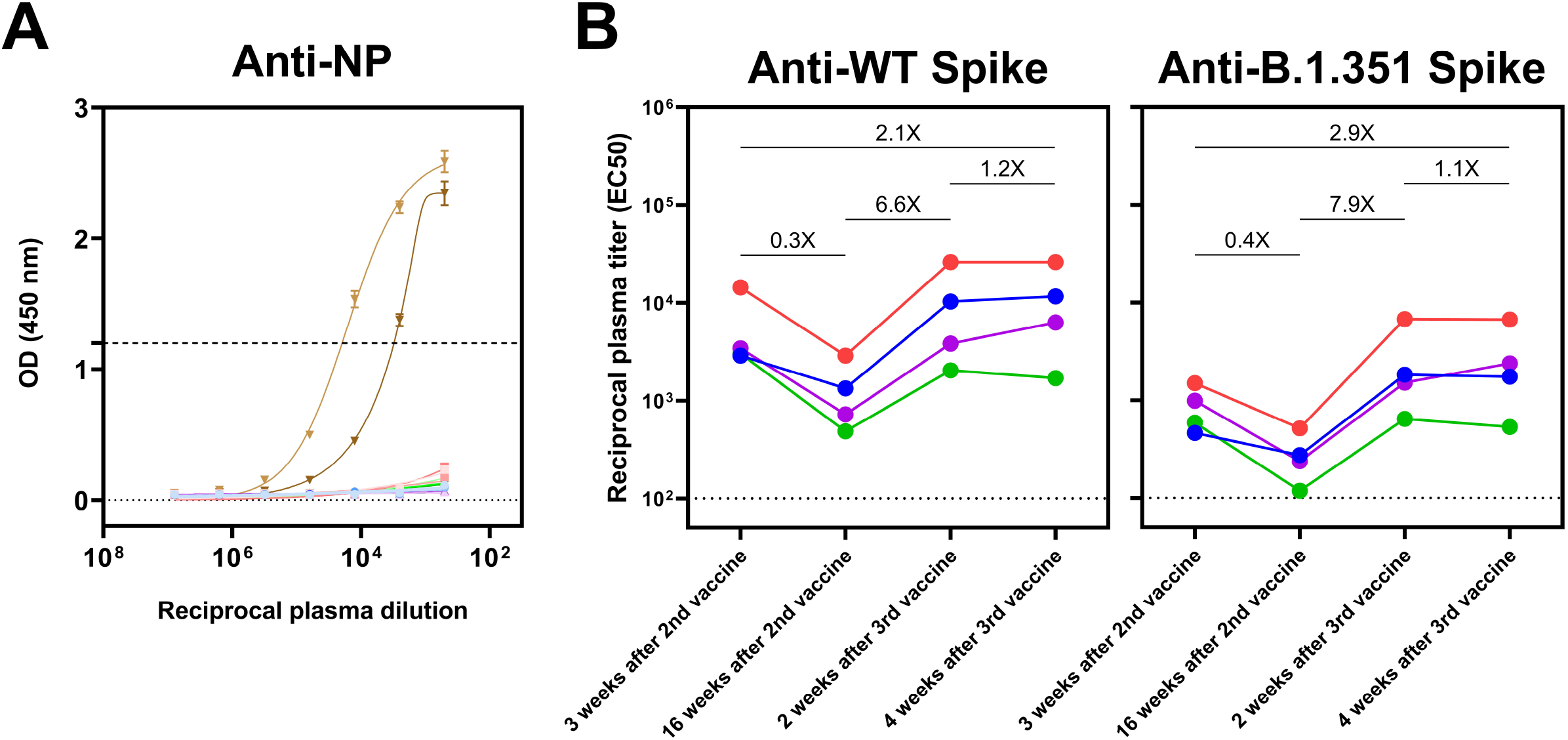
Binding profiles of individual samples against SARS-CoV-2 nucleoprotein (NP) and spike. A, Two negative control plasma from healthy pre-SARS-CoV-2 patients and two positive control plasma from convalescent SARS-CoV-2 patients were included in the experiments. Data are shown as mean ± SEM of two technical replicates. Colors denote the samples from individual vaccinees at each timepoint: blue = Vaccinee #1, red = Vaccinee #2, purple = Vaccinee #3, green = Vaccinee #4. Negative control plasma are denoted in light gray and gray and positive control plasma are denoted in light brown and brown. B, Plasma samples collected at the indicated timepoints were tested for binding to non-variant (WT) and B.1.351 SARS-CoV-2 spike. The average fold change in reciprocal plasma titer (EC_50_) between two timepoints are denoted. Colors denote individual vaccinees: blue = Vaccinee #1, red = Vaccinee #2, purple = Vaccinee #3, green = Vaccinee #4.

**Figure 2.**
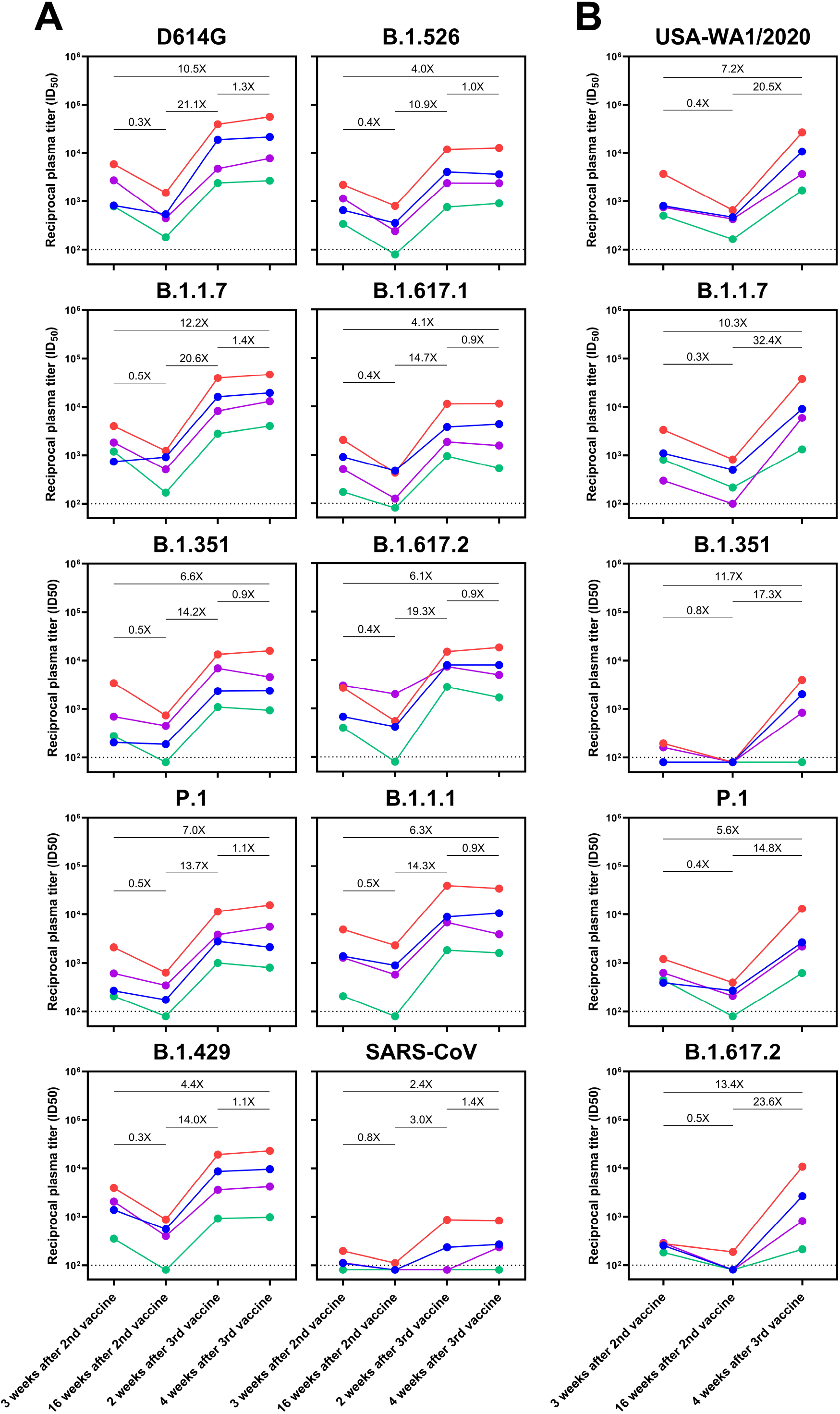
Immunogenicity of three SARS-CoV-2 vaccinations in healthy individuals. A, Plasma samples were tested for neutralizing capability against recombinant vesicular stomatitis virus (rVSV) pseudotyped with spike from non-variant SARS-CoV-2 with D614G mutation, SARS-CoV-2 variants, or SARS-CoV. B, Plasma samples were tested for neutralizing capability against authentic non-variant SARS-CoV-2 (USA-WA1/2020) and SARS-CoV-2 variants in a cytopathic effect reduction assay. In all panels, average fold change in reciprocal plasma titer (ID_50_) between two timepoints are denoted. The limit of detection in both neutralization assays is ID_50_ = 100, and samples below the limit of detection are arbitrarily shown as 80. Colors denote individual vaccinees: blue = Vaccinee #1, red = Vaccinee #2, purple = Vaccinee #3, green = Vaccinee #4.

In this observational study examining the antibody response of individuals receiving a third vaccine dose, a robust boost in the strength of neutralizing antibodies, as well as breadth against SARS-CoV-2 variants was observed in all individuals. Although our cohort size is small, the similarity of results across individuals indicates a common effect. As observed in other studies, each of the individuals had a decay of neutralizing antibodies over time (3, 4), and demonstrated reduced neutralizing titer against some of the SARS-CoV-2 variants (5-7). This combination of temporal decay of antibody titer and the emergence of SARS-CoV-2 variants may therefore lead to a loss of protection in some vaccinated individuals. We demonstrate herein that a third COVID-19 vaccine dose strongly boosts neutralizing antibody titers, including against rapidly spreading variants such as B.1.617.2 (delta variant) (**Figure 2**), reaching levels beyond that induced by two vaccinations. Importantly, this heterologous three-vaccine regimen had similar trends as a previously reported homologous three-vaccine schedule (12), and may therefore serve as one practical option. While it remains to be seen whether a third booster dose is needed, we illustrate here that should it be required, such a strategy strongly bolsters antibody responses and could therefore be another instrument in the arsenal for full control of this pandemic.

## Materials and Methods

This study was reviewed and approved by the Institutional Review Board of Columbia University. The four individuals in this study received their first dose of BNT162b2 vaccine in December 2020, second dose of BNT162b2 vaccine in January 2021 (three weeks after the initial dose), and a dose of Ad26.COV2.S vaccine in May 2021 (four months after the second vaccination). Plasma was collected three weeks after the second vaccination, 16 weeks after the second vaccination, two weeks after the third vaccination, and four weeks after the third vaccination. Enzyme-linked immunosorbent assay (ELISA) to quantify anti-SARS-CoV-2 spike and nucleoprotein antibodies, recombinant vesicular stomatitis virus-based SARS-CoV-2 pseudovirus neutralization assay, and authentic SARS-CoV-2 neutralization assay, were performed as previously described (15).

## Data Availability

All relevant data are included with the manuscript.

## Acknowledgements

We are grateful to the participants in this study for contributing to SARS-CoV-2 research. There was no applicable funding for this study.

